# Cost-Effectiveness Analysis of the mRNA-1345 RSV Vaccine for Adults in the United States: Clinical and Economic Value

**DOI:** 10.1101/2025.08.01.25332812

**Authors:** Katherine Hicks, Ziyi Xiao, Kelly Fust, Michele Kohli, Kyle Paret, Weijie Zhang, Maria de Pilar Martin Matos, Keya Joshi, Parinaz Ghaswalla

**Affiliations:** RTI Health Solutions, Durham, NC, USA; Quadrant Health Economics, Cambridge, ON, Canada; Moderna Inc, Cambridge, MA, USA

**Keywords:** Cost-effectiveness, respiratory syncytial virus, vaccines, mRNA, economic value, clinical value

## Abstract

**Background:** The clinical and economic burden of respiratory syncytial virus (RSV) is significant; millions of patients seek medical care due to RSV infections each year. Recently approved vaccines offer an important opportunity to reduce this burden in populations that are at high risk due to certain medical conditions or older age. This study evaluates the public health impact and cost-effectiveness of vaccination with mRNA-1345 against RSV in adults.

**Methods:** A static decision-analytic model was developed to estimate the public health impact and cost-effectiveness of vaccinating high-risk adults aged 18-59 years and all adults aged ≥ 60 years in the 5-year period following vaccination against RSV with a single dose of mRNA-1345 in the United States (US). RSV vaccination rates in the analysis were assumed to be equal to 2019-2020 influenza season vaccination rates. Vaccine effectiveness against RSV-related acute respiratory disease and RSV-associated lower respiratory tract infection over time was modeled using nonlinear waning curves that reflect clinical trial data.

**Results:** According to the model, vaccinating the high-risk adult US population aged 18-59 years and all US adults aged ≥ 60 years with mRNA-1345 would avert over 280,000 hospitalizations, 19,000 deaths, and result in about 244,000 quality-adjusted life-years gained compared with an unvaccinated population over a 5-year period. Vaccination costs approximately $23.2 billion and reduces direct and indirect costs by $10.8 billion and $13.3 billion, respectively, resulting in net savings of more than $810 million from a societal perspective, making vaccination with mRNA-1345 in this population cost saving.

**Conclusions:** Vaccination with mRNA-1345 is a cost-saving strategy for the prevention of RSV in high-risk and older US adults and has the potential for substantial public health impact.

**KEY SUMMARY POINTS:** *Why carry out this study? (1-2 bullets):*

- The clinical and economic burden of respiratory syncytial virus (RSV) in high-risk and older adults is significant
- The mRNA-1345 RSV vaccine has been approved in adults aged ≥ 60 years in 2024 and high-risk adults 18-59 years old in 2025, but the potential clinical benefit and cost-effectiveness of RSV vaccination has not been established.

*What did the study ask?/What was the hypothesis of the study? (1 bullet):*

- The study objective was to examine the public health impact and cost-effectiveness of RSV vaccination with mRNA-1345 compared with no vaccination in high-risk adults aged 18-59 years and all adults aged ≥ 60 years in the United States (US) in the 5-year period following vaccination.

*What were the study outcomes/conclusions? (1 bullet point):*

- Vaccination with mRNA-1345 results in 280,000 fewer hospitalizations, over 19,000 fewer deaths, and over $810 million in economic savings from a societal perspective compared with not vaccinating against RSV over a 5-year period.

*What has been learned from the study? (1-2 bullets):*

- These analyses suggest the mRNA-1345 vaccine is a cost-saving strategy for the prevention of RSV in US adults across age groups and has the potential for substantial public health impact.
- Our findings may inform clinical and policy decision-making regarding the value of RSV vaccination in adults and support efforts to expand access among younger high-risk populations.

**PLAIN LANGUAGE SUMMARY:** Respiratory syncytial virus (RSV) is a common virus that causes lung infections. It can be especially dangerous for older adults and people with health conditions like asthma, heart disease, or diabetes. Each year, RSV causes many hospital stays and even deaths in the United States.

A new vaccine called mRNA-1345 was recently approved to protect adults aged 60 and older, and high-risk adults aged 18-59. This study looked at whether giving the vaccine to these groups would help prevent RSV infections and save money.

Researchers used a mathematical model to compare what might happen over 5 years with and without the vaccine. They found that the vaccine could prevent over 280,000 hospital stays and more than 19,000 deaths. Even though the vaccine costs money, it would save more than $800 million by reducing medical bills and helping people stay healthy and working.

RSV can place a heavy burden on individuals and the healthcare system. This study shows that the mRNA-1345 vaccine could protect the most vulnerable adults while also being a valuable investment for society.

## INTRODUCTION

Respiratory syncytial virus (RSV) is a common respiratory virus that affects an estimated 64 million people and results in 160,000 deaths per year worldwide, with the highest mortality rates in high-income countries occurring among adults aged 65 years and older [1, 2]. Common symptoms of RSV infection include nasal congestion, cough, shortness of breath, fever, and fatigue, making it difficult to differentiate RSV infections from influenza or COVID-19 [3]. Many RSV infections resolve without the need for medical attention in young, healthy individuals, but RSV infections can be dangerous and even fatal in older adults and adults with significant comorbidities [4].

Increasing age is an independent risk factor for severe outcomes in adults. In addition, certain underlying conditions—such as asthma, chronic obstructive pulmonary disease (COPD), coronary artery disease (CAD), congestive heart failure (CHF), chronic kidney disease (CKD), chronic liver disease, diabetes mellitus, or severe obesity (body mass index [BMI] ≥ 40)—can also increase the risk [5]. Patients with 1 or more of these conditions are also considered at “high risk” for severe disease.

The clinical and economic burden of RSV is significant; millions of patients seek medical care due to RSV infections each year [6]. During the 2024-2025 RSV season in the United States (US), as many as 350,000 adults were hospitalized with RSV, and up to 23,000 died, according to the Centers for Disease Control and Prevention (CDC) [6]. While older adults (≥ 60 years old) are at highest risk of severe disease, the RSV medical burden is significant across all adult age groups [7]. Carrico et al. [8] estimated RSV cases in US adults aged 50-59 years cost $1.1 billion annually from a societal perspective; they also estimated a high economic burden due to RSV for adults aged 18-59 years with high-risk conditions for RSV. Averin and colleagues [9] estimated that, while adults aged over 60 years account for 50% of the economic burden of RSV in adults, adults aged 18-59 years at high risk for severe RSV additionally account for 27% of the total economic burden of RSV, with costs of $25.0 billion ($15.2 billion in direct costs and $9.7 billion in indirect costs).

Since 2023, several vaccines have been approved by the US Food and Drug Administration (FDA) for the prevention of RSV-related lower respiratory tract disease (RSV-LRTD), defined as RSV with at least 2 lower respiratory signs or symptoms, in adults aged over 60 years [10], including RSVPreF (Abrysvo) [11], RSVPreF3 (Arexvy) [12], and mRNA-1345 (mRESVIA) [13]. In light of the availability of these vaccines, the CDC’s Advisory Committee on Immunization Practices (ACIP) recommends that all adults over 75 years of age and adults aged 50 years and older at high risk for severe RSV receive a single dose of an RSV vaccine [5, 14, 15]. As of April 2025, the CDC estimated 38.1% of high-risk US adults aged 60-74 years and 47.5% of US adults aged ≥75 years had received the RSV vaccine [16]. Because these vaccines have become available only in the past few years, their impact on the clinical and economic burden of RSV infection in older adults has not been established. In 2025, the approval for mRNA-1345 as expanded to adults aged 18-59 years who are at increased risk for LRTD caused by RSV [17]. In this study, we developed a decision-analytic model designed to estimate the potential public health impact and cost-effectiveness of vaccination with a single dose of mRNA-1345 compared with no vaccine in US high-risk adults aged 18-59 years and all adults aged ≥ 60 years (hereafter, “the indicated population”), consistent with the labeled indication for mRNA-1345 [17].

## METHODS

### Model Overview

A static decision-analytic model was developed to estimate the public health impact and cost-effectiveness of vaccination of the indicated population in the 5-year period following vaccination with mRNA-1345 in the US [18]. The model estimated cases of RSV-related acute respiratory disease (RSV-ARD) and RSV-LRTD, quality-adjusted life-years (QALYs), clinical outcomes to estimate public health burden, and economic outcomes from a societal perspective to estimate cost-effectiveness. Patients who developed RSV-ARD were classified as having RSV-LRTD or having RSV-No LRTD (without at least 2 lower respiratory signs or symptoms). Cost-effectiveness was measured as the incremental cost-effectiveness ratio (ICER) calculated as the incremental cost per QALY saved from the societal perspective. Figure 1 presents the decision tree applied annually in the model to calculate RSV outcomes.

**Figure 1.**
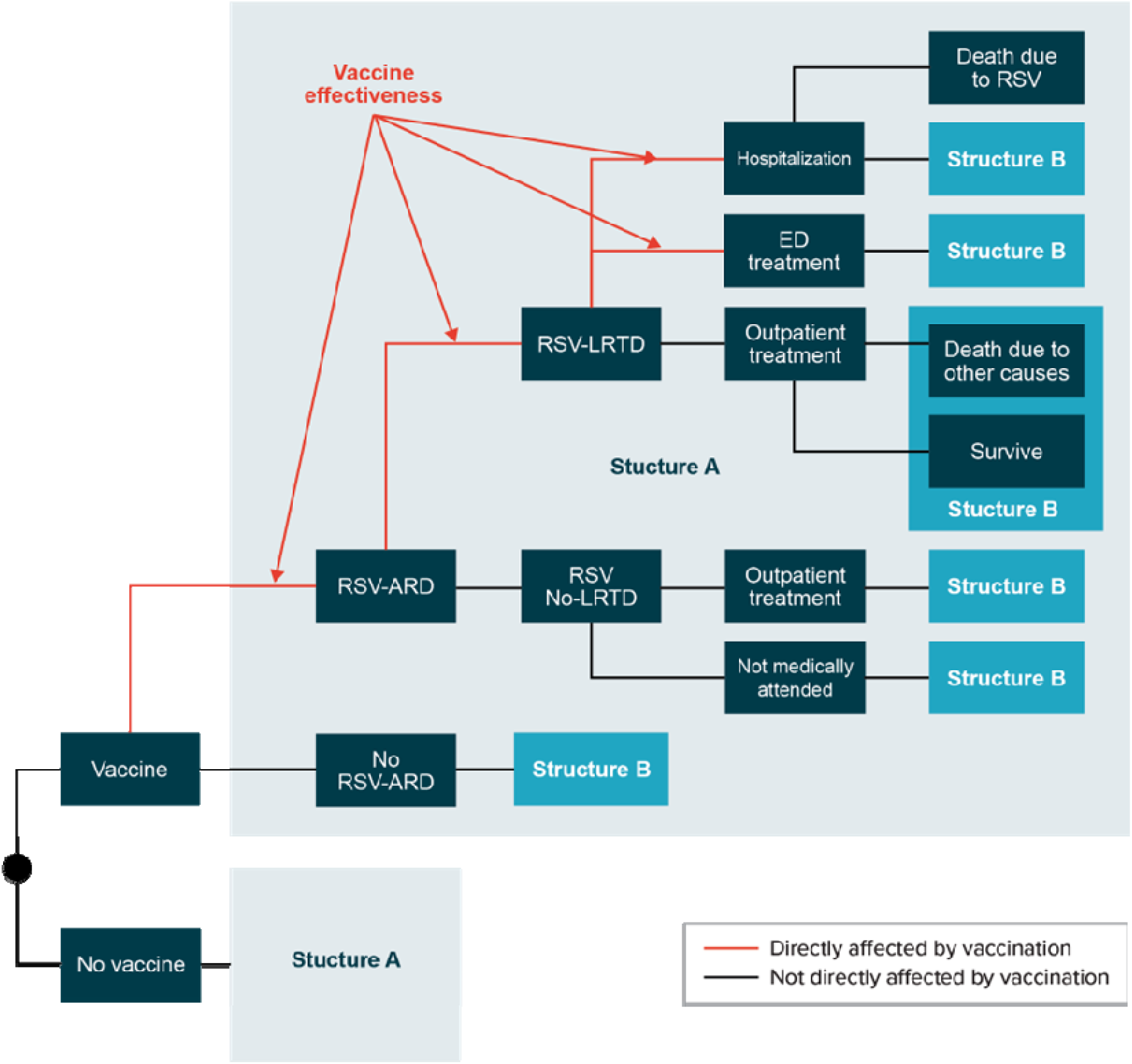
Model Structure. ED = emergency department; ICER = incremental cost-effectiveness ratio; RSV = respiratory syncytial virus; RSV-ARD = RSV-related acute respiratory disease; RSV-LRTD = RSV-related lower respiratory tract disease.

### Population

The population for the base-case analysis was high-risk adults aged 18-59 years and all adults aged ≥ 60 years, in line with the FDA approval for mRNA RSV vaccination [14, 17, 19]. Consistent with the ACIP’s recommendations, for the purposes of model analyses, “high risk” was defined as having at least 1 of the following conditions: asthma, COPD, CAD, CHF, CKD, chronic liver disease, diabetes mellitus, or severe obesity (BMI ≥ 40) [14].

### Intervention

In this study, vaccination with mRNA-1345 is compared with no vaccine for the observed population. The mRNA-1345 vaccine was assumed to be effective against RSV-ARD and RSV-LRTD, which reflected clinical trial data; the vaccine effectiveness for hospitalization and emergency department (ED) treatment was assumed to be the same as the clinical trial effectiveness against RSV-LRTD [13]. Therefore, in this analysis, preventing a case of RSV-ARD or RSV-LRTD was also assumed to prevent all RSV-related downstream health consequences (e.g., hospitalizations and death).

### Model Inputs

Table 1 presents key model inputs and sources. All model inputs and sources are listed in Table S1.

**Table 1.**
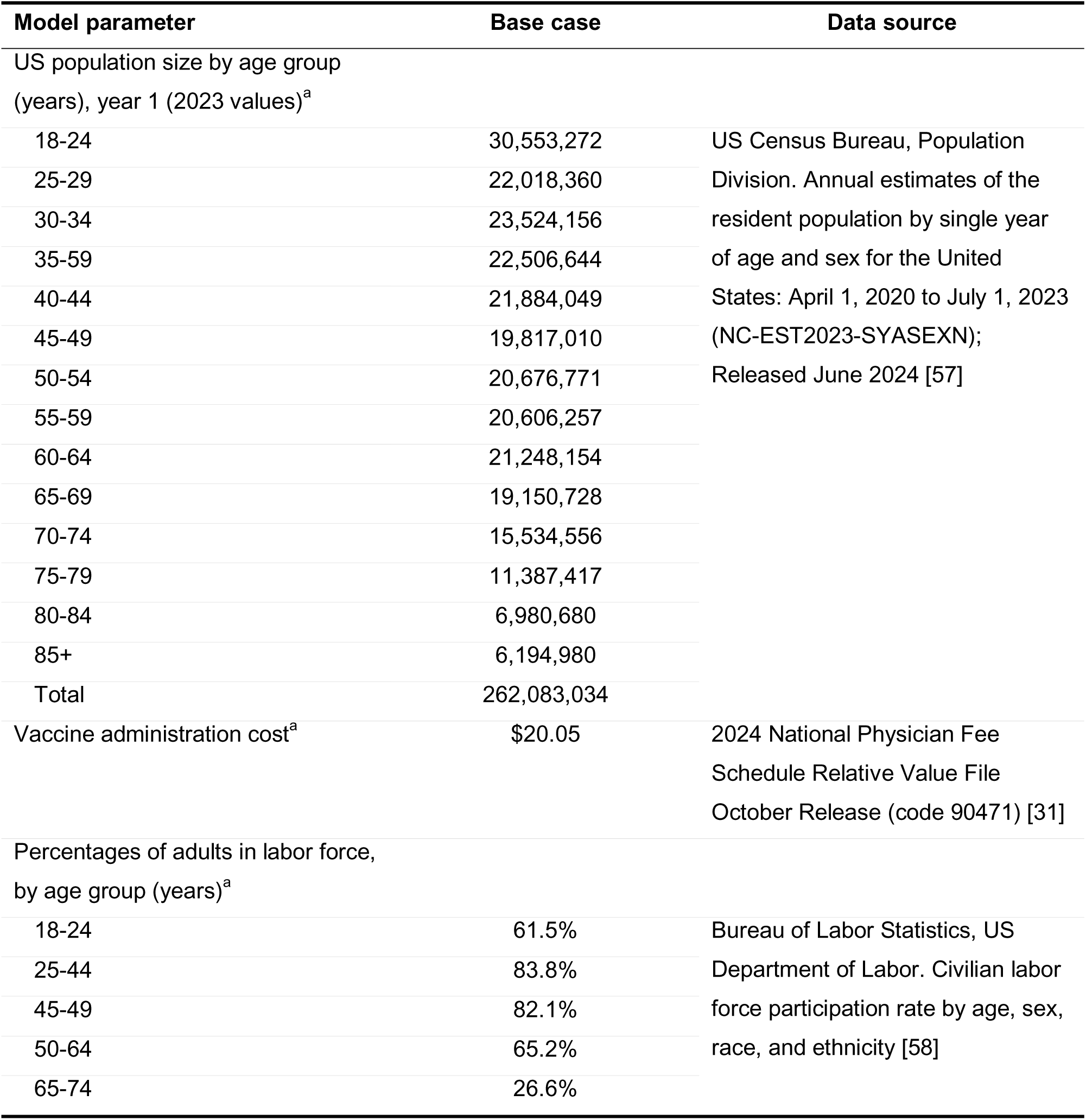
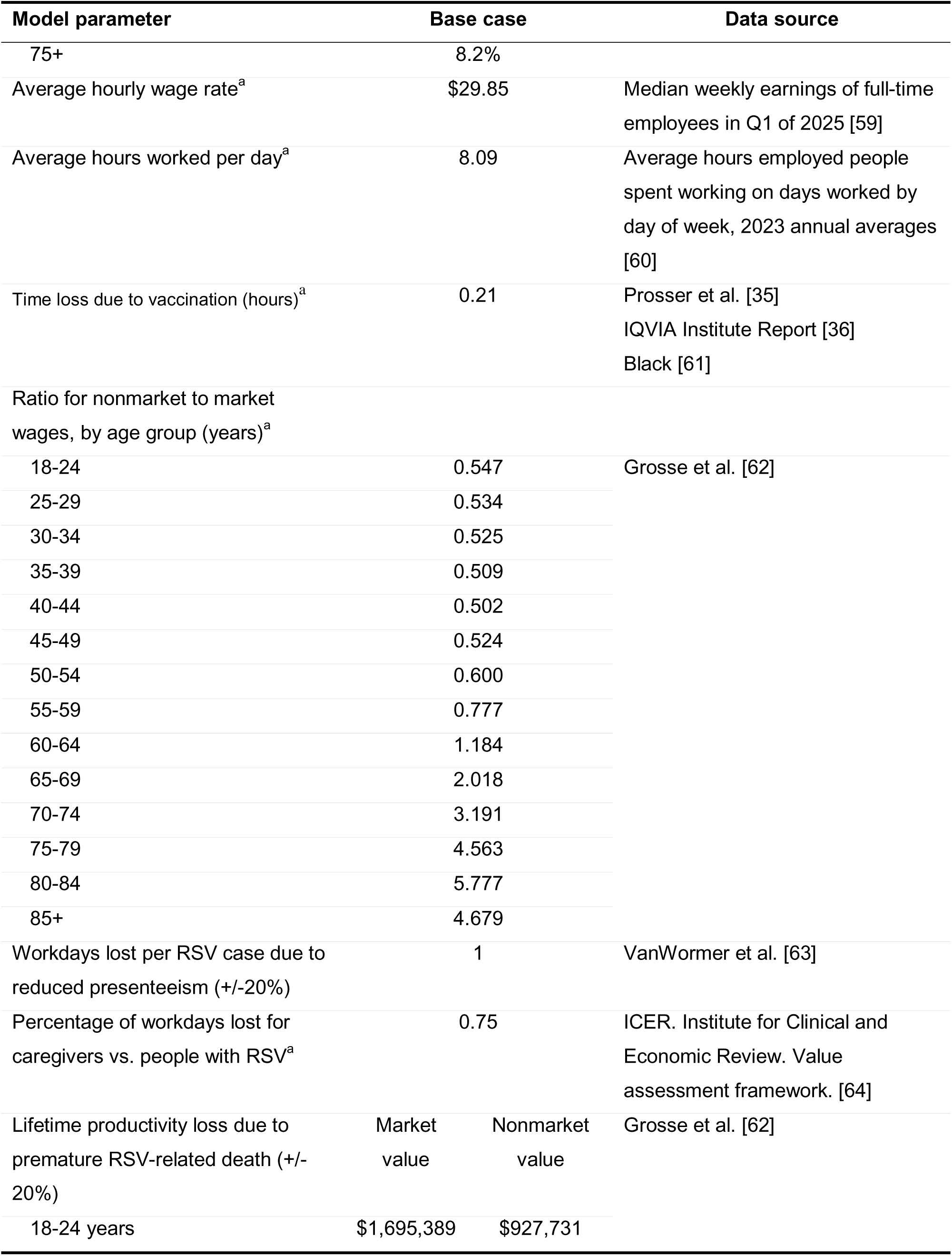
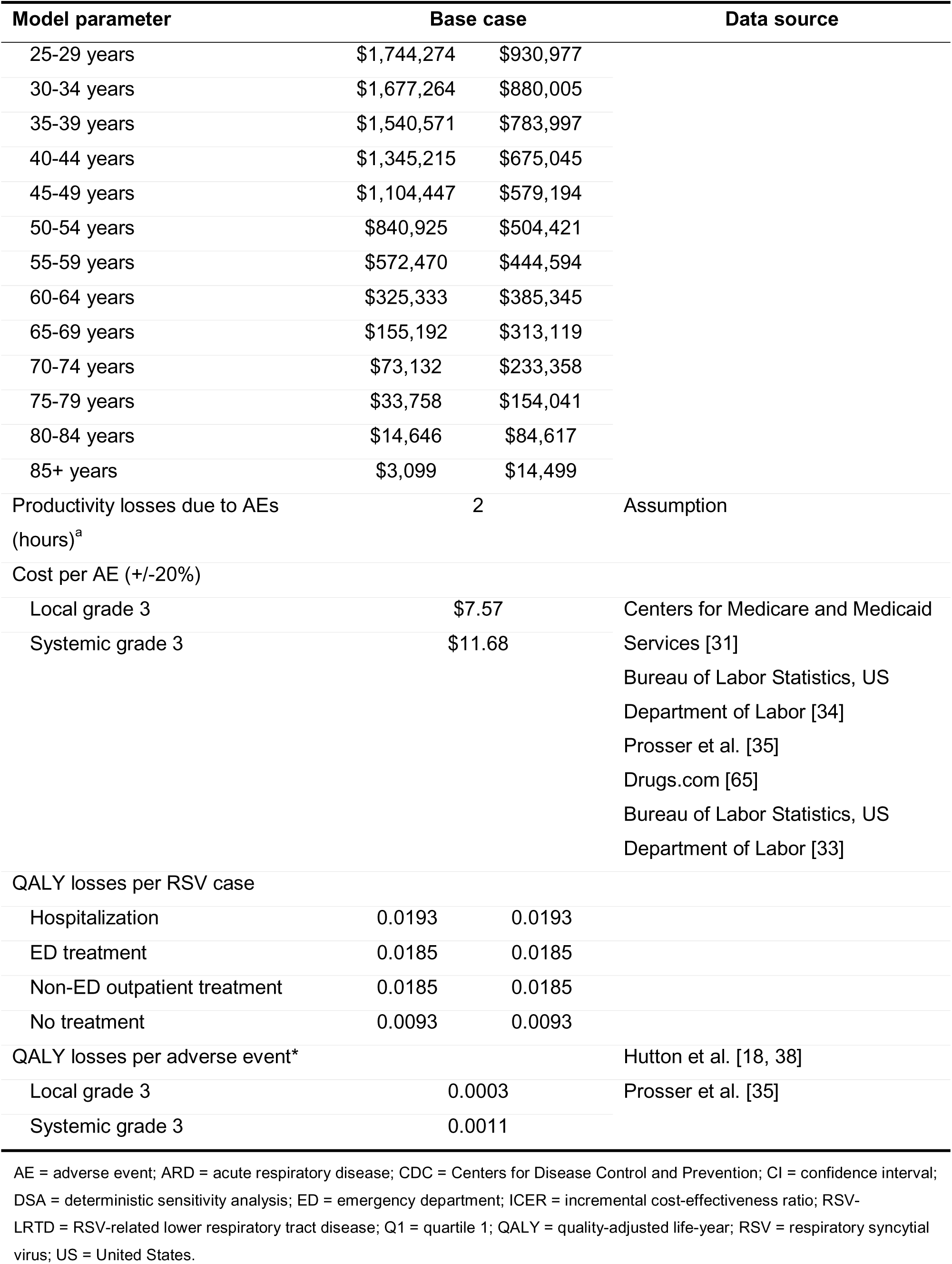
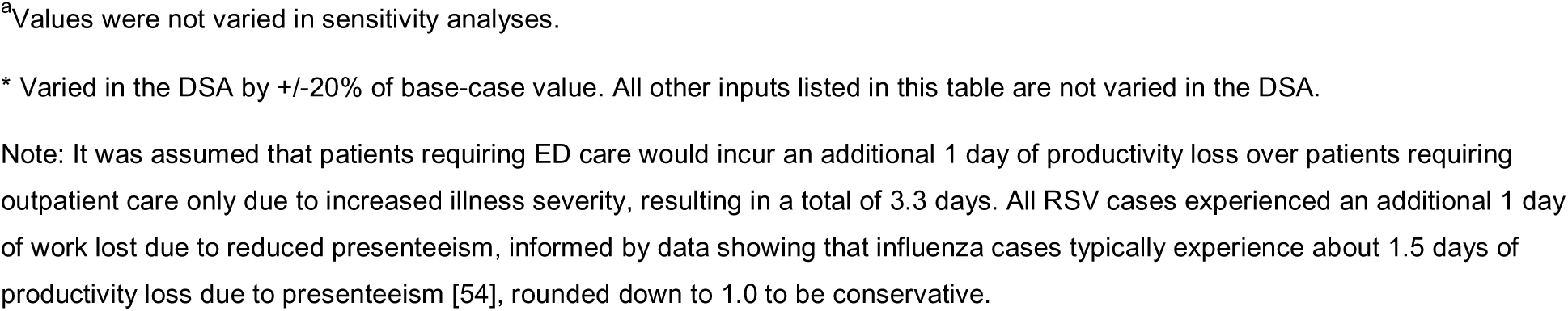
Selected Model Inputs: Population Size and Unvaccinated Transition Probabilities (Target Population—All Adults Aged ≥ 60 Years + Adults Aged 18-59 Years and at High Risk for Severe RSV)

| Model parameter | Base case | Data source |
| --- | --- | --- |
| US population size by age group (years), year 1 (2023 values)<sup>a</sup> |  |  |
| 18-24 | 30,553,272 | US Census Bureau, Population Division. Annual estimates of the resident population by single year of age and sex for the United States: April 1, 2020 to July 1, 2023 (NC-EST2023-SYASEXN); Released June 2024 [57] |
| 25-29 | 22,018,360 |  |
| 30-34 | 23,524,156 |  |
| 35-59 | 22,506,644 |  |
| 40-44 | 21,884,049 |  |
| 45-49 | 19,817,010 |  |
| 50-54 | 20,676,771 |  |
| 55-59 | 20,606,257 |  |
| 60-64 | 21,248,154 |  |
| 65-69 | 19,150,728 |  |
| 70-74 | 15,534,556 |  |
| 75-79 | 11,387,417 |  |
| 80-84 | 6,980,680 |  |
| 85+ | 6,194,980 |  |
| Total | 262,083,034 |  |
| Vaccine administration cost<sup>a</sup> | \$20.05 | 2024 National Physician Fee Schedule Relative Value File October Release (code 90471) [31] |
| Percentages of adults in labor force, by age group (years)<sup>a</sup> |  |  |
| 18-24 | 61.5% | Bureau of Labor Statistics, US Department of Labor. Civilian labor force participation rate by age, sex, race, and ethnicity [58] |
| 25-44 | 83.8% |  |
| 45-49 | 82.1% |  |
| 50-64 | 65.2% |  |
| 65-74 | 26.6% |  |

| Model parameter | Base case |  | Data source |
| --- | --- | --- | --- |
| 75+ | 8.2% |  |  |
| Average hourly wage rate<sup>a</sup> | \$29.85 | | Median weekly earnings of full-time employees in Q1 of 2025 [59] |
| Average hours worked per day<sup>a</sup> | 8.09 |  | Average hours employed people spent working on days worked by day of week, 2023 annual averages [60] |
| Time loss due to vaccination (hours)<sup>a</sup> | 0.21 |  | Prosser et al. [35]<br>IQVIA Institute Report [36]<br>Black [61] |
| Ratio for nonmarket to market wages, by age group (years)<sup>a</sup> |  |  |  |
| 18-24 | 0.547 |  | Grosse et al. [62] |
| 25-29 | 0.534 |  |  |
| 30-34 | 0.525 |  |  |
| 35-39 | 0.509 |  |  |
| 40-44 | 0.502 |  |  |
| 45-49 | 0.524 |  |  |
| 50-54 | 0.600 |  |  |
| 55-59 | 0.777 |  |  |
| 60-64 | 1.184 |  |  |
| 65-69 | 2.018 |  |  |
| 70-74 | 3.191 |  |  |
| 75-79 | 4.563 |  |  |
| 80-84 | 5.777 |  |  |
| 85+ | 4.679 |  |  |
| Workdays lost per RSV case due to reduced presenteeism (+/-20%) | 1 |  | VanWormer et al. [63] |
| Percentage of workdays lost for caregivers vs. people with RSV<sup>a</sup> | 0.75 |  | ICER. Institute for Clinical and Economic Review. Value assessment framework. [64] |
| Lifetime productivity loss due to premature RSV-related death (+/- 20%) | Market value | Nonmarket value | Grosse et al. [62] |
| 18-24 years | \$1,695,389 | \$927,731 | |
| 25-29 years | \$1,744,274 | \$930,977 | |
| 30-34 years | \$1,677,264 | \$880,005 | |
| 35-39 years | \$1,540,571 | \$783,997 | |
| 40-44 years | \$1,345,215 | \$675,045 | |
| 45-49 years | \$1,104,447 | \$579,194 | |
| 50-54 years | \$840,925 | \$504,421 | |
| 55-59 years | \$572,470 | \$444,594 | |
| 60-64 years | \$325,333 | \$385,345 | |
| 65-69 years | \$155,192 | \$313,119 | |
| 70-74 years | \$73,132 | \$233,358 | |
| 75-79 years | \$33,758 | \$154,041 | |
| 80-84 years | \$14,646 | \$84,617 | |
| 85+ years | \$3,099 | \$14,499 | |
| Productivity losses due to AEs (hours)<sup>a</sup> | 2 |  | Assumption |
| Cost per AE (+/-20%) |  |  |  |
| Local grade 3 | \$7.57 | | Centers for Medicare and Medicaid Services [31]<br>Bureau of Labor Statistics, US Department of Labor [34]<br>Prosser et al. [35]<br>Drugs.com [65]<br>Bureau of Labor Statistics, US Department of Labor [33] |
| Systemic grade 3 | \$11.68 | | |
| QALY losses per RSV case |  |  |  |
| Hospitalization | 0.0193 | 0.0193 |  |
| ED treatment | 0.0185 | 0.0185 |  |
| Non-ED outpatient treatment | 0.0185 | 0.0185 |  |
| No treatment | 0.0093 | 0.0093 |  |
| QALY losses per adverse event* |  |  | Hutton et al. [18, 38] |
| Local grade 3 | 0.0003 |  | Prosser et al. [35] |
| Systemic grade 3 | 0.0011 |  |  |

AE = adverse event; ARD = acute respiratory disease; CDC = Centers for Disease Control and Prevention; CI = confidence interval; DSA = deterministic sensitivity analysis; ED = emergency department; ICER = incremental cost-effectiveness ratio; RSV-LRTD = RSV-related lower respiratory tract disease; Q1 = quartile 1; QALY = quality-adjusted life-year; RSV = respiratory syncytial virus; US = United States.
<sup>a</sup>Values were not varied in sensitivity analyses.
\* Varied in the DSA by +/-20% of base-case value. All other inputs listed in this table are not varied in the DSA.
Note: It was assumed that patients requiring ED care would incur an additional 1 day of productivity loss over patients requiring outpatient care only due to increased illness severity, resulting in a total of 3.3 days. All RSV cases experienced an additional 1 day of work lost due to reduced presenteeism, informed by data showing that influenza cases typically experience about 1.5 days of productivity loss due to presenteeism [54], rounded down to 1.0 to be conservative.

#### RSV-Related Incidence

The incidence of RSV-ARD infection without vaccination was derived from Falsey et al. [20], who prospectively evaluated 3 cohorts of patients in Rochester, New York, including a cohort of “high-risk” patients, defined as those 21 years or older with CHF or COPD. Using methods similar to those described in Averin et al. [9], data from McLaughlin et al. [21] and Weycker [22] were used to estimate RSV-related hospitalization, ED visits, and outpatient rates per 100,000 people by age group and risk (high or not high for adults aged ≥ 60 years; adults aged 18-59 years in the indicated population were all high risk) [21, 22]. An adjustment factor of 1.5 was applied to the calculated rates to account for underreporting of RSV due to polymerase chain reaction test sensitivity. A calibration process was then used to estimate the percentage of RSV-ARD cases that result in LRTD, percentage of RSV-LRTD cases that require hospitalization and ED visits (versus outpatient care only), and the percentage of RSV-No LRTD cases without LRTD that require outpatient (versus no outpatient) care, by risk level and age group, so that the inputs of interest most closely reflected the available incidence data and most closely aligned with clinically relevant logic requiring that (1) within an age group, non–high-risk people did not have worse outcomes than high-risk people and (2) within a risk level, people in younger age groups did not have worse outcomes than people in older age groups. It was assumed that all LRTD cases receive medical attention, while patients with RSV-No LRTD do not experience hospitalization or ED visits.

#### Vaccination Rates and Effectiveness

Since limited or no RSV vaccination coverage rates for the observed populations are available, RSV vaccination rates in the analysis were assumed to be equal to 2019-2020 influenza season vaccination rates [23]. In the base case, vaccine effectiveness against RSV-ARD and RSV-LRTD over time was modeled using nonlinear waning curves (Figure 2). A nonlinear waning profile was assumed to reflect clinical trial data showing that RSV neutralizing antibody titers remained robust and well above baseline levels between 12 and 24 months post-vaccination, supported by durable cell-mediated immunity across age and risk groups and by emerging evidence linking antibody levels with protection [24–26]. The nonlinear models were fitted to monthly trial data through a median follow-up of 18.8 months to estimate the duration of protection following vaccination with mRNA-1345. The model used the logarithm of time as the independent variable and monthly log incidence rate ratio as the dependent variable. As shown in Figure 2, modeled vaccine effectiveness against RSV-LRTD and RSV-ARD was projected up to 60 months, with the rate of waning slowing beyond 24 months. Consistent with recommendations from the ISPOR-SMDM Modeling Good Research Practices Task Force [27], the base-case analyses used a 5-year time horizon to capture the full benefits of vaccination. Annual effectiveness measures were then calculated, weighted by incidence seasonality patterns and assuming that RSV vaccination would be given in September, prior to the typical start of the RSV season in the US. The same incidence pattern was applied each year, recognizing that historically, the seasonal patterns have varied across years.

**Figure 2.**
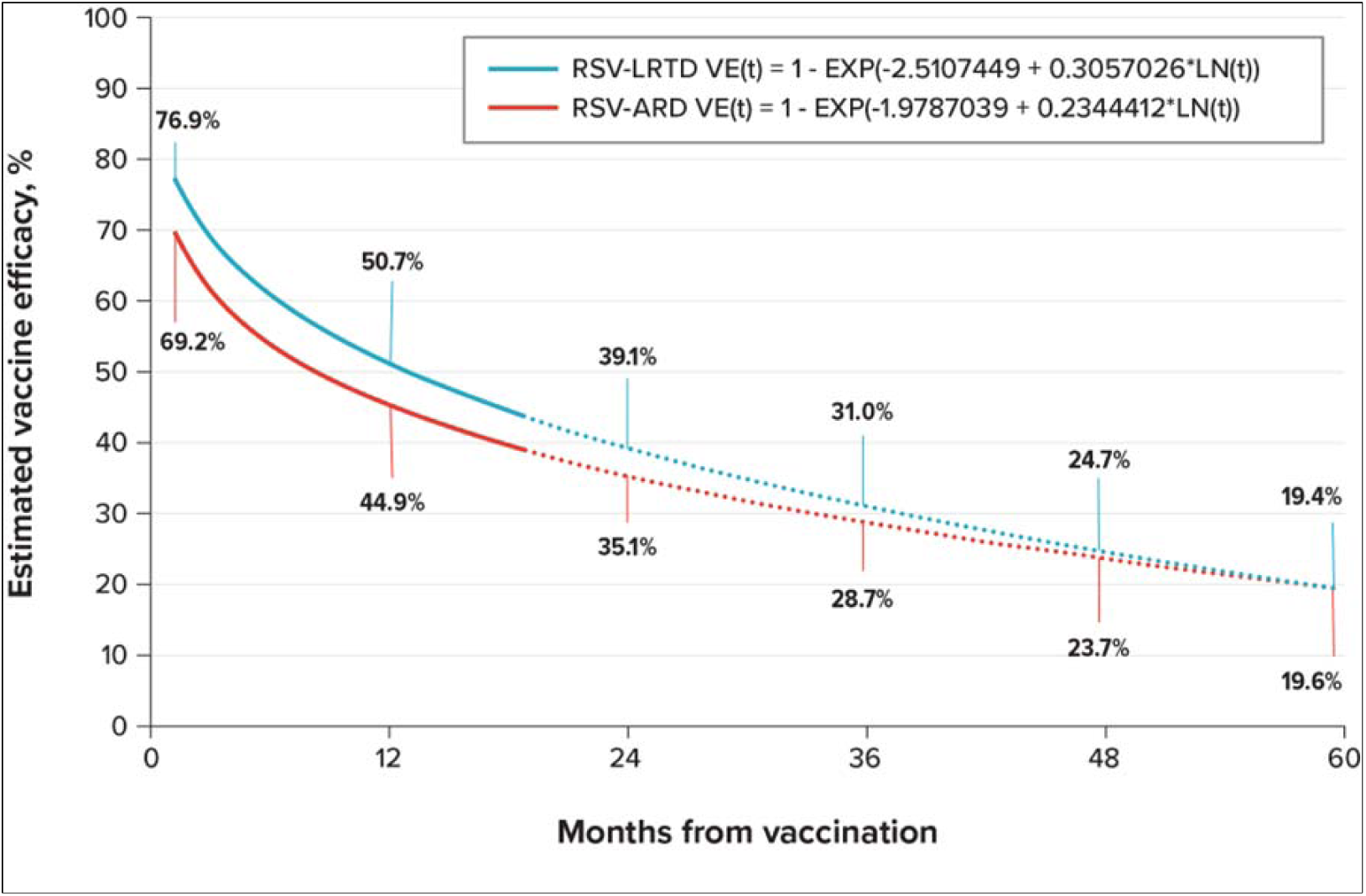
Projected Vaccine Effectiveness Over 5 Years Using Nonlinear Waning Assumptions Applied to Vaccine Effectiveness: Base-Case Analysis. RSV = respiratory syncytial virus; RSV-ARD = RSV-related acute respiratory disease; RSV-LRTD = RSV-related lower respiratory tract disease; VE = vaccine effectiveness. Note: Solid lines indicate clinical trial data, and dashed lines indicate extrapolated data.

#### Costs

The costs applied in the model for hospitalized and outpatient RSV cases were estimated from data reported by Wyffels and colleagues [28] as the difference between the preindex and postindex costs for hospitalized and outpatient cases, respectively, each as weighted averages from costs of high-risk and non–high-risk patients [29]. Since Wyffels et al. [28] did not report ED costs, estimations by Averin et al. [9] for patients immunocompetent with chronic medical conditions were used to estimate costs for RSV cases requiring ED visits from all-cause healthcare expenditures during the 1-year period from the beginning of an RSV-LRTD episode. All costs were inflated to 2024 US dollars using the medical care component of the US Consumer Price Index (CPI) [29]. A cost of $0 was assumed for patients who did not require any medical treatment.

For the base-case analysis, the unit cost of the mRNA-1345 vaccine was aligned with the current list prices of available RSV vaccines [30]. The administration cost was estimated from the 2024 Centers for Medicare and Medicaid Services National Physician Fee Schedule [31] using the Healthcare Common Procedure Coding System (HCPCS) code 90471 for immunization administration.

Nonhealthcare costs included the loss of productive time from missed work due to receiving the mRNA-1345 vaccine or for RSV cases. The cost of lost productivity was calculated using data from the US Bureau of Labor Statistics by multiplying the percentage of adults in the labor force by mean hourly income for the total population and the average number of hours worked per day, then further multiplying by the days expected to be lost from work due to either receiving the vaccination or RSV infection [32–34]. Productivity losses due to absenteeism and presenteeism for RSV cases were included. In the base-case analysis, premature mortality–related productivity losses were included and estimated using the human capital method; both market and nonmarket productivity were incorporated [35, 36]. Lost work productivity for caregivers was included for hospitalized cases.

#### QALYs

QALY losses were incurred due to RSV morbidity and mortality and vaccination adverse events. Age-specific population utility values were used to calculate the QALYs lost due to early death from RSV, discounted to present value. Values were calculated as a weighted average of Short-Form 6-Dimension (SF-6D) health state scores for males and females from Hanmer and Kaplan [37]. The values provided by Hutton [18, 38] were used as the base-case estimates for the number of QALYs lost by patients requiring hospitalization, ED, and outpatient care, assuming that those seeking ED care would experience QALY losses equivalent to those seeking outpatient treatment. Consistent with another recent cost-effectiveness analysis of RSV vaccination [39], it was assumed that patients who did not seek medical care would experience 50% of the QALY losses of patients receiving outpatient care. No QALY losses were assumed for caregivers. QALY losses for vaccine adverse events were assumed to be the same as those applied in Prosser et al. [35].

### Outcomes

Clinical outcomes for estimating public health burden included the number of hospitalizations, ED visits, outpatient visits, QALYs lost, and deaths due to RSV. Economic outcomes for estimating cost-effectiveness included RSV-related direct medical costs, vaccine-related costs (including unit cost, administration, and adverse events), indirect costs (including productivity loss due to RSV-related morbidity and mortality, productivity loss due to vaccination and adverse events, caregiver productivity loss), and total costs from a societal perspective. ICERs from mRNA-1345 versus no vaccination were also estimated.

### Scenario and Sensitivity Analyses

Five additional sets of scenario analyses were conducted to examine ICERs under different assumptions: (1) populations with varying age and risk categories (high-risk 18- to 49-year-olds, high-risk 50- to 59-year-olds, high-risk 60- to 74-year-olds, all adults aged ≥ 75 years, and all adults aged ≥ 60 years), (2) a healthcare perspective, (3) caregiver impact excluded, (4) linear waning of vaccine effectiveness (1.9% and 1.8% monthly reduction for RSV-LRTD and RSV-ARD, respectively) applied over 2-year and 3-year time horizons, and (5) individuals aged 18-59 years with each of the individual risk conditions indicating high risk for severe RSV (asthma, COPD, CAD, CHF, CKD, chronic liver disease, diabetes mellitus, and severe obesity).

A deterministic sensitivity analysis (DSA) and probabilistic sensitivity analysis (PSA) using 10,000 iterations of the model were also conducted. DSA input value variations are detailed in Tables S-2. Specific ranges around base-case values of interest were observed in the DSA. Otherwise, ranges reflecting 20% variation from the base-case values were used. Variance data around the base-case values were used as ranges for inputs in the PSA, where available. For all other inputs, a user-specified percentage variation from a base value was used to represent a 95% confidence interval and the standard error corresponding to that interval was calculated and used to define the distribution. The percentage variation was 10% or 20%, depending on the assumed level of uncertainty for the PSA. Ten percent was used for input values if the point estimates were directly estimated from data sources; otherwise, 20% was used.

### Ethics Statement

Data for this study were collected and analyzed from published research and did not require additional ethical review.

## RESULTS

### Base-Case Analysis

Assuming similar vaccination coverage to influenza in the 2019-2020 season, approximately 75 million US adults out of a total eligible population of approximately 135 million would be vaccinated with mRNA-1345. Without vaccination, the model predicted over 0.23 million hospitalizations, almost 0.24 million ED visits, approximately 2.8 million outpatient visits, almost 15,000 deaths attributable to RSV, and over 16.8 million QALYs lost on average per year in the observed population. Over a 5-year time horizon, vaccination with a single dose of mRNA-1345 prevented 280,000 hospitalizations (25% reduction), over 263,000 ED visits, over 2.8 million outpatient visits, over 19,000 deaths (26% reduction), and 244,000 QALYs lost compared with no vaccination (Table 2). Vaccination with mRNA-1345 in the indicated population was estimated to cost over $23 billion, including vaccine administration costs, acquisition costs, and costs related to adverse events. However, when direct medical and indirect costs related to RSV infection were considered, vaccination resulted in savings of more than $810 million compared with no vaccination (Table 2). Vaccination of the indicated population with mRNA-1345 is not only clinically beneficial but also cost saving.

**Table 2.** Base-Case^a^ Analysis Results (5-year period)

|  | mRNA-1345 | No vaccination | Difference<sup>▮b</sup> |
| --- | --- | --- | --- |
| Clinical outcomes (thousands of cases) |  |  |  |
| RSV-ARD cases | 32,011 | 39,856 | -7,846 |
| RSV-LRTD cases | 4,843 | 6,210 | -1,367 |
| Hospitalizations | 862 | 1,144 | -282 |
| ED visits | 932 | 1,195 | -263 |
| Outpatient visits | 11,121 | 13,969 | -2,848 |
| Deaths | 55 | 74 | -19 |
| QALYs lost | 83,848 | 84,092 | -243 |
| Economic outcomes (\$ billions) | | | |
| Vaccination costs | \$23.2 | \$0.0 | \$23.2 |
| Direct RSV medical costs | \$37.9 | \$48.7 | -\$10.8 |
| Indirect RSV costs | \$52.0 | \$65.3 | -\$13.3 |
| Total costs<sup>▮c</sup> | \$113.2 | \$114.0 | -\$0.8 |
| ICER |  |  | Cost saving |

ED = emergency department; ICER = incremental cost-effectiveness ratio; QALY = quality-adjusted life-year; RSV = respiratory syncytial virus; RSV-ARD = RSV-related acute respiratory disease; RSV-LRTD = RSV-related lower respiratory tract disease.
<sup>a</sup>Base-case scenario considers the high-risk population aged 18-59 years and all people aged ≥60 years, the societal perspective, impact on caregivers, and nonlinear vaccine effectiveness waning.
<sup>b</sup> mRNA-1345 vaccine compared with no vaccine.
<sup>▮c</sup> Total of all direct costs, vaccination costs, and indirect costs.

### Scenario Analyses

ICERs from the scenario analyses are listed in Table 3; additional outcomes are presented in Tables S-3 through S-6. Across the observed age subgroups, the ICER was just over $18,000 per QALY gained for vaccinating high-risk 18- to 49-year-old adults with mRNA-1345 compared with no vaccination but, similar to the base case, was cost saving for 50- to 59-year-old and 60- to 74-year-old high-risk subgroups. When considering all risk levels, the ICER per QALY gained was cost saving for vaccination with mRNA-1345 compared with not vaccinating in the ≥60-year age group and was over $11,000 per QALY gained in the ≥75-year age group.

**Table 3.** Scenario Analysis Results.

| **Scenario<sup>a</sup>** | **ICERs** |
| --- | --- |
| Base-case scenario<sup>a</sup> | Cost saving |
| Subgroup defined by age and RSV risk level |  |
| High risk, 18-49 years old | \$18,027 |
| High risk, 50-59 years old | Cost saving |
| High risk, 60-74 years old | Cost saving |
| All 75+ | \$11,714 |
| All 60+ | Cost saving |
| Healthcare perspective | \$51,200 |
| No caregiver impact | Cost saving |
| Vaccine effectiveness scenario |  |
| Monthly linear waning (1.9% for RSV-LRTD and 1.8% for RSV-ARD) over 3-year time horizon | \$22,467 |
| Monthly linear waning over 2-year time horizon | \$46,535 |
| Subgroup aged 18-59 years with conditions indicating high risk for severe RSV |  |
| Asthma | \$43,418 |
| Diabetes | Cost saving |
| CAD | Cost saving |
| CKD | Cost saving |
| Chronic liver disease | Cost saving |
| COPD | Cost saving |
| CHF | Cost saving |
| Morbid obesity | Cost saving |

CAD= coronary artery disease, CHF=congestive heart failure; CKD= chronic kidney disease; COPD=chronic obstructive pulmonary disease; RSV-ARD = RSV-related acute respiratory disease; RSV-LRTD = RSV-related lower respiratory tract disease; RSV = respiratory syncytial virus.
<sup>a</sup>Base-case scenario considers the high-risk population aged 18-59 years and all people aged ≥60 years, the societal perspective, impact on caregivers, and nonlinear vaccine effectiveness waning

Excluding caregiver impact also resulted in cost savings. When considering the healthcare perspective, the ICER was $51,200 per QALY gained for mRNA-145 vaccination compared with no vaccination. Assuming vaccine effectiveness wanes linearly monthly over 3- and 2-year time horizons, the ICERs were $22,467 and $46,535 per QALY gained for mRNA-1345 compared with no vaccination, respectively.

When evaluating the impact of vaccination in 18- to 59-year-old people with specific conditions, indicating high risk for severe RSV, mRNA-1345 improved all observed clinical outcomes for all conditions and was cost saving in populations with all conditions except asthma ($43,418 per QALY gained). Clinical gains were highest in people with asthma, diabetes, and morbid obesity. The only positive ICER was observed in population aged 18-59 years with asthma ($43,418 per QALY gained). Clinical outcomes, including the number of hospitalizations, outpatient visits, and deaths, were universally improved with vaccination compared with no vaccination across all scenarios.

### Sensitivity Analyses

Results of the DSA are presented in Table 4. Model results were most sensitive to costs of RSV cases requiring outpatient treatment, percentage experiencing RSV-ARD, and percentage of hospitalized RSV cases resulting in RSV-related death. Observed ICERs were between cost saving and $21,352..

**Table 4.** Deterministic Sensitivity Analysis Results.

| Model input | ICER: mRNA-1345 vs No vaccine |  |
| --- | --- | --- |
|  | Lower bound | Upper bound |
| Costs of RSV cases requiring outpatient treatment | \$21,352 | Cost saving |
| Percentage experiencing RSV-ARD | \$21,079 | Cost saving |
| Percentage of hospitalized RSV cases resulting in RSV-related death | \$10,037 | Cost saving |
| Percentage of RSV cases experiencing LRTD<sup>a</sup> | \$9,090 | Cost saving |
| Percentage of RSV-LRTD patients requiring hospitalization<sup>a</sup> | \$6,475 | Cost saving |
| Lifetime productivity loss due to premature RSV-related death | \$3,015 | Cost saving |
| Percentage of RSV-No LRTD patients requiring outpatient treatment<sup>a</sup> | \$547 | Cost saving |
| Costs of RSV-LRTD cases requiring hospitalization | \$80 | Cost saving |
| Percentage of RSV-LRTD patients requiring ED department treatment<sup>a</sup> | Cost saving | Cost saving |
| Costs of RSV cases requiring ED treatment | Cost saving | Cost saving |
| Percentage of vaccinations with mRNA-1345 experiencing local grade 3 AEs | Cost saving | Cost saving |
| Adverse Events with mRNA-1345: Local Grade 3 Cost | Cost saving | Cost saving |
| Adverse Events with mRNA-1345: Local Grade 3 QALY lost | Cost saving | Cost saving |
| Adverse Events with mRNA-1345: Systemic Grade 3 Cost | Cost saving | Cost saving |
| Adverse Events with mRNA-1345: Systemic Grade 3 QALY lost | Cost saving | Cost saving |
| QALY loss per RSV case (for all levels of care) | Cost saving | Cost saving |

ED = emergency department; ICER = incremental cost-effectiveness ratio; RSV = respiratory syncytial virus; RSV-ARD = RSV-related acute respiratory disease; RSV-LRTD = RSV-related lower respiratory tract disease.
<sup>a</sup> Input parameters for the percentage of RSV cases experiencing RSV-LRTD, percentage of RSV-LRTD patients requiring hospitalization, percentage of RSV-No LRTD patients requiring outpatient treatment, and percentage of RSV-LRTD patients requiring an ED visit were derived via calibration to reflect incidence rates from McLaughlin et al. [66]; relative rates of hospitalization, ED, and outpatient visits due to LRTD for high-risk versus non-high-risk populations from Weycker et al. [22]; and 1.5 adjustment for underdetection [21].

Results from the PSAs are presented in Figure 3. Figure 3i demonstrated that vaccination with mRNA-1345 in the observed populations dominated the no-vaccine alternative across all willingness-to-pay thresholds. Figure 3ii demonstrates that vaccinating the indicated population with mRNA-1345 was more effective and cost saving (dominant) compared with no vaccination in over 50% of model iterations.

**Figure 3.**
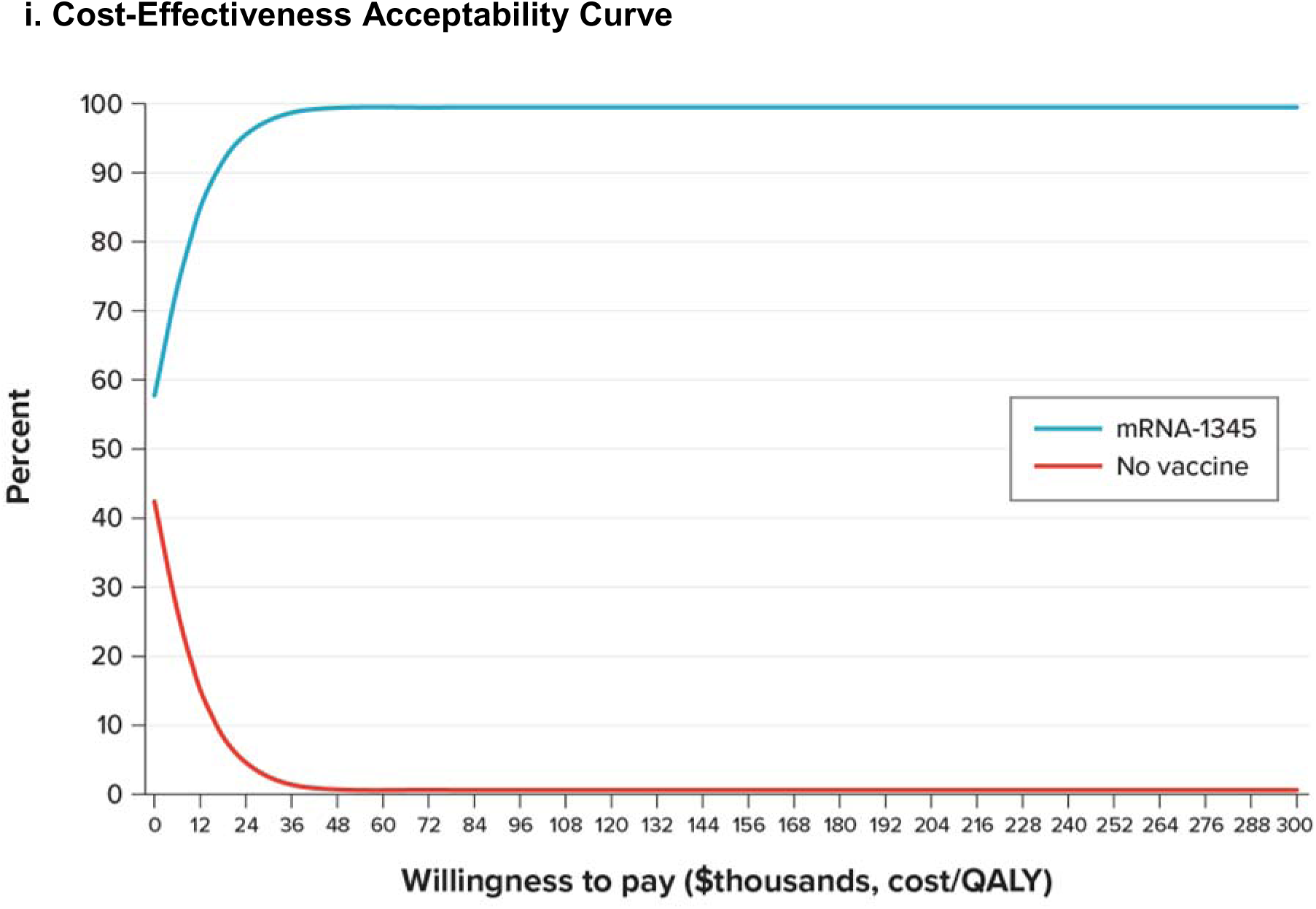
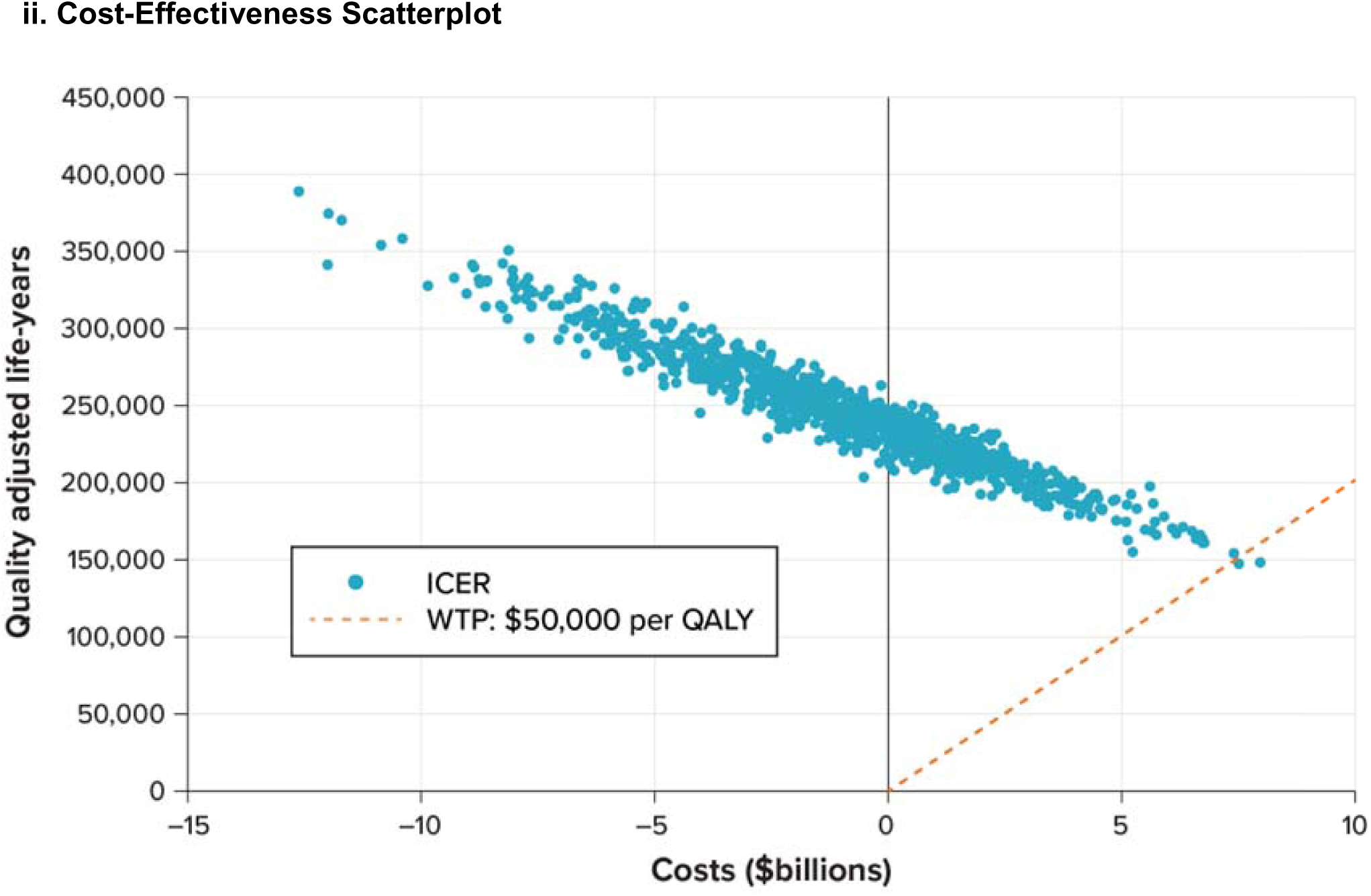
Probabilistic Sensitivity Analysis. CEAC = cost-effectiveness acceptability curve; ICER = incremental cost-effectiveness ratio; QALY = quality-adjusted life-year; WTP = willingness-to-pay threshold.

## DISCUSSION

At vaccination rates comparable to those for influenza, RSV vaccination with mRNA-1345 produces significant clinical and economic benefit across all indicated age and risk groups. In the base-case scenario, vaccinating the indicated population resulted in about 244,000 fewer lost QALYs, 282,000 fewer hospitalizations, 263,000 fewer ED visits, and 19,000 fewer deaths over a 5-year period than if there were no vaccination. mRNA-1345 vaccination in this population also yielded over $810 million in savings overall. Scenario analyses and sensitivity analyses demonstrated that cost savings of RSV vaccination were maintained across many variations in input values. When conducting condition-specific analyses among the 18- to 59-year-old high-risk population, the clinical and economic benefits of vaccination across individual risk conditions was greatest for groups with asthma, diabetes, and morbid obesity.

To date, 5 economic evaluations of RSV vaccination in US older adults have been published [38, 40–43]. Two of these evaluations occurred prior to the availability of RSV vaccines for adults in the US [42, 43]. In 2000, Gessner et al. [42] found that a potential RSV vaccine would be cost-effective among a US cohort of 65-year-olds, with costs per QALY equal to $5,342. In 2022, Herring et al. [43] performed a value-based pricing analysis with 2 base-case analyses (hypothetical vaccine vs. no vaccine) and found that RSV vaccination among US adults ≥ 60 years is likely to be cost-effective at price levels similar to those of other vaccines recommended for older adults in the US. The other 3 studies were conducted in 2 protein subunit RSV vaccines (Arexvy [GSK] and Abrysvo [Pfizer]) following their approval by the FDA and their recommendation by ACIP in 2023 for adults aged ≥ 60 years [44, 45]. In 2024, an economic evaluation of RSV vaccination compared with no vaccination in older adults (≥ 65 years) was funded by the CDC and led by Hutton et al. [38], who found that if vaccination uptake was 20% among older adults, outpatient RSV illness could be reduced by 10.2%-11.1%, hospitalizations and hospitalized death reduced by 13.5%-14.7%, savings in healthcare costs were $460-$500 million, savings in productivity costs were $400-$450 million, and 15,700-18,000 QALYs were saved. In 2024, La et al. [41] conducted a cost-effectiveness analysis on the adjuvanted RSVpreF3 vaccine among older US adults (≥ 60 years) and found that vaccination resulted in an ICER of $18,430 per QALY gained and an incremental cost savings of $4.5 billion over 5 years compared with no vaccination [41]. Li et al. [40] conducted a multicohort analysis in 2025 among older European adults (≥ 60 years) to identify key drivers of cost-effectiveness, reporting that vaccine price, hospitalization costs, inpatient fatality ratio, age, and vaccine duration of protection were all influential.

These previous cost-effectiveness studies on RSV vaccination have focused on older adults (≥ 60 years) only. To our knowledge, this is the first cost-effectiveness analysis of RSV vaccination in the scientific literature that includes high-risk adults aged 18-59 years, though the clinical and economic burdens of RSV in this population have been increasingly recognized [46, 47]. The results presented here align with previously published work reporting that RSV vaccination among older adults is cost-effective at $50,000 and $150,000 willingness-to-pay thresholds [41]; furthermore, results of the scenario analyses expanded that knowledge to show that also including RSV vaccination in younger high-risk adults aged 18-59 years—while not cost saving as an independent subgroup—maintained cost savings when combined with adults aged 60 years and older in the base-case scenario.

As with all decision-analytic studies, this analysis is subject to several limitations, particularly related to data availability. Several studies suggest that RSV infections are underdetected; for example, McLaughlin et al. [21] suggest that detection was 1.5 times greater when a second specimen type was added to polymerase chain reaction testing, and Rozenbaum et al. [48] also notes that studies using *International Classification of Diseases, Tenth Revision* (ICD-10) discharge diagnosis codes to detect RSV infection substantially underestimate the true burden of the disease. Similarly, Howa et al. [49] found that the hospitalization burden of RSV in adults is underestimated, in part due to a lack of routine testing. We applied a 1.5 adjustment factor to estimated RSV cases in our analysis, consistent with the study by McLaughlin and colleagues [21], but this was still an estimate, and other similar analyses have reported up to 2.2 times higher rates of underreporting [50]. Furthermore, vaccine efficiency was assumed to impact hospitalization and LRTD rates similarly; however, evidence suggests vaccines reduce severe outcomes like hospitalization much more, and therefore, our analysis may be conservative relative to the real-world vaccine effectiveness [21, 51]. Studies of nonmedically attended RSV are also limited, further leading to an underestimation of the full burden of RSV. While the study by Falsey et al. [20] was carefully designed to allow identification of symptomatic RSV cases in the community and among hospitalized cases, there was a limit to the size of the cohort that could be followed prospectively. Given the small sample size, there is uncertainty associated with the estimates of incidence. The high-risk population is extremely heterogeneous, including many people who have multiple conditions resulting in high risk of severe RSV. Therefore, representing the high-risk population overall with 1 set of model input values presents multiple challenges and requires significant generalization. The most fitting estimates identified in the scientific literature were applied for that generalized group in this analysis. Furthermore, RSV incidence varies year to year, and the estimate of vaccine economic value depends in part on the severity of the season, as seen in influenza [52, 53]. Wyffels et al. [28] defined the outpatient cohort as those who were not hospitalized within 1 day of RSV diagnosis. While the 180-day postindex period used in that study may have allowed for follow-up hospitalization costs, there may be some mixture of patients with acute infection who were treated in outpatient or hospital settings. Given the data available, it was also assumed in this study that individuals acquired 1 infection per season; however, if individuals acquired more than 1 infection per season and both are prevented by the vaccine, the value of the vaccine may also increase.

There are few data on the productivity loss associated with reduced presenteeism due to RSV [38], and assumptions have been made on the basis of a previously published influenza model [54]. Previous reviews have concluded that there is a large degree of heterogeneity in time loss due to influenza because of both variation in study design and the economic behavior of individuals [55, 56].

If RSV vaccination coverage rates can be increased to influenza vaccination levels, there could be a substantial reduction of the public health burden of RSV infections among US older and high-risk younger adults. Under the current ACIP recommendation for RSV vaccines (all adults aged ≥ 75 years and high-risk adults aged 50-74 years) [5, 14, 15], mRNA-1345 could prevent over 252,000 RSV-related hospitalizations and over 18,000 RSV-related deaths over a 5-year period. Expanding RSV vaccination recommendations to include all populations indicated for mRNA-1345 (high-risk adults aged 18-59 years and all adults aged ≥60 years) would generate an even larger public health benefit while saving overall costs versus no vaccine.

## CONCLUSION

These analyses suggest that vaccination with mRNA-1345 is a cost-saving strategy for the prevention of RSV in high-risk US adults aged 18-59 years and all adults aged ≥60 years and has the potential for substantial public health impact in these populations. Our findings may inform clinical and policy decision-making regarding the value of RSV vaccination in adults and support efforts to expand access among younger high-risk populations.

## ACKNOWLEDGEMENTS

Medical writing was provided by Taylor Tibbs and Sara Musetti Jenkins; editing was provided by Arthur Iannacone and John Forbes; and graphic design was provided by Jason Crouch. All acknowledged individuals are employees of RTI Health Solutions, who received funding for manuscript preparation from Moderna, Inc.

## FUNDING

Funding for this study and for medical writing and editing support was provided by Moderna, Inc. as part of a research contract with RTI Health Solutions.

## CONFLICTS OF INTEREST

This study was conducted by RTI Health Solutions under the direction of Moderna and was funded by Moderna, Inc. Maria de Pilar Martin Matos, Keya Joshi, and Parinaz Ghaswalla are employees of Moderna, Inc. Katherine Hicks, Ziyi Xiao, and Kyle Paret are employees of RTI Health Solutions. Kelly Fust is a consultant to Quadrant Health Economics, Inc. Michele Kohli is a shareholder in Quadrant Health Economics, Inc. This study was conducted by RTI Health Solutions under the direction of Moderna and was funded by Moderna, Inc.

## AUTHOR CONTRIBUTIONS

All authors contributed to the study conception and design. Material preparation, data collection, and analysis were performed by Ziyi Xiao and Katherine Hicks. The first draft of the manuscript was written with medical writing support, and all authors commented on previous versions of the manuscript. All authors read and approved the final manuscript.

## DATA AVAILABILITY

Data sources for this study are publicly available, as described in Table 1.

## Notes

### Author Declarations

Originally located at published resources. Published resource includes reference 13 as noted in lines 145-148 in the manuscript

