## Supplementary Material for "Cost-Effectiveness Analysis of the mRNA-1345 RSV Vaccine for Adults in the United States: Clinical and Economic Value"

Moderna Inc,

Cambridge, MA, USA

Table S-1 Additional Population Size and Unvaccinated Transition Probabilities (Target Population All Adults Aged ≥ 60 Years + 18-59 High Risk)

| Model parameter | Value (DSA range) | | | | | | | Data source |
| --- | --- | --- | --- | --- | --- | --- | --- | --- |
| Annual incidence of RSV-ARD, unvaccinated | | | | | | | | |
| % with symptomatic RSV-ARD (+/-20%) | | | | | | | | |
| 18-59 | 6.70 | | | | | | | Falsey et al. (2005) [1] |
| 60+ | 5.73 | | | | | | | Derived from Falsey et al. (2005) [1] and data on file at Moderna |
| % of RSV-ARD patients with RSV-LRTD by age group (years), unvaccinated (+/-20%) ^‡^ | | | | | | | | |
| 18-59 | 15.20 | | | | | | | Estimated by calibration using McLaughlin (2022) [2] (McLaughlin JM 2022), Weycker (2024) [3] |
| 60-64 | 13.45 | | | | | | |  |
| 65-74 | 14.47 | | | | | | |  |
| 75-84 | 19.30 | | | | | | |  |
| 85+ | 24.65 | | | | | | |  |
| % of RSV-LRTD patients requiring each level of care by age group (years), unvaccinated (+/-20%) | | | | | | | | |
| Care setting: | Hospital | ED | | Outpatient | | No treatment | | Estimated by calibration using McLaughlin (2022) [2], Weycker (2024) [3] |
| 18-49 | 7.12 | 20.69 | | 72.19 | | 0 | |  |
| 50-59 | 21.40 | 17.37 | | 61.23 | | 0 | |  |
| 60-64 | 13.75 | 14.80 | | 71.45 | | 0 | |  |
| 65-74 | 25.39 | 20.09 | | 54.52 | | 0 | |  |
| 75-84 | 28.89 | 20.07 | | 51.04 | | 0 | |  |
| 85+ | 31.16 | 20.02 | | 48.82 | | 0 | |  |
| % of RSV-No LRTD patients requiring each level of care by age group (years) (+/-20%) | | | | | | | | |
| Care setting: | Hospital | ED | | Outpatient | | No treatment | | Estimated by calibration using McLaughlin (2022) [2], Weycker (2024) [3] |
| 18-49 | 0 | 0 | | 25.3 | | 74.7 | |  |
| 50-59 | 0 | 0 | | 29.9 | | 70.1 | |  |
| 60-64 | 0 | 0 | | 24.07 | | 75.93 | |  |
| 65-74 | 0 | 0 | | 33.44 | | 66.56 | |  |
| 75-84 | 0 | 0 | | 39.61 | | 60.39 | |  |
| 85+ | 0 | 0 | | 47.30 | | 52.70 | |  |
| % of hospitalized RSV cases resulting in RSV-related death, by age group (+/-20%) | | | | | | | | |
| 18-44 | 2.4 (1.9-6.0) | | | | | | | Pastula et al. (2017) [4], Branche et al. (2023) [5], Havers et al. (2024) [6] |
| 45-59 | 4.3 (3.1-6.0) | | | | | | |  |
| 60-64 | 7.6 (3.9-7.6) | | | | | | | Falsey et al. (2005) [1], Hutton et al. (2024) Appendix [7], Tseng et al. 2020 [8] |
| 65-74 | 7.6 (4.3-7.6) | | | | | | |  |
| 75+ | 7.6 (5.7-9.4) | | | | | | |  |
| Death due to other causes* | Age-specific | | | | | | | General population mortality from 2018 US life tables. Arias et al. (2020) [9] |
| Percentage of eligible individuals vaccinated with mRNA-1345, by age group (if considering mRNA-1345)* | | | | | | | | 2019-2020 influenza vaccination rates from CDC [10] |
| 18-49 | 38.4% | | | | | | |  |
| 50-59 | 50.6% | | | | | | |  |
| 60+ | 69.8% | | | | | | |  |
| Vaccine unit cost* | $290 | | | | | | | Data provided by Moderna based on current market price of available RSV vaccines |
| Vaccine administration cost* | $20.05 | | | | | | | 2024 National Physician Fee Schedule Relative Value File October Release (code 90471) [11] |
| Direct costs per RSV case, by age group and care setting | | | | | | | | |
| Care setting: | Hospital | | ED | | Outpatient | | No treatment | Hospital: Wyffels et al. (2020) [12]  ED: Averin et al. (204) [13]  Outpatient: Wyffels et al. (2020) [12] |
| 18-49 | $12,198  ($9,123-$48,962) | | $3,258  ($2,334-6,074) | | $2,334  ($128-$2,334) | | $0 |  |
| 50-59 | $12,198  ($9,123-$48,962) | | $4,776  ($2,334-6,074) | | $2,334  ($128-$2,334) | | $0 |  |
| 60-64 | $12,198  ($9,123-$48,962) | | $4,776  ($2,334-6,074) | | $2,334  ($128-$2,334) | | $0 |  |
| 65-74 | $12,198  ($9,123-$48,962) | | $4,776  ($2,334-6,074) | | $2,334  ($128-$2,334) | | $0 |  |
| 75-84 | $12,198  ($9,123-$48,962) | | $4,149  ($2,334-6,074) | | $2,334  ($128-$2,334) | | $0 |  |
| 85+ | $12,198  ($9,123-$48,962) | | $4,379  ($2,334-6,074) | | $2,334  ($128-$2,334) | | $0 |  |
| Workdays lost per RSV case due to absenteeism, by age group and care setting* | | | | | | | | |
| Care setting: | Hospital | | ED | | Outpatient | | No treatment | Hospital: Ackerson et al. (2020) [14]  ED: Assumption (1 additional day of productivity loss relative to outpatient cases to reflect increased severity of illness)  Outpatient / No treatment: Falsey et al. (2005) [1], Chit et al. (2015) [15] |
| 18-49 | 7.53 | | 3.3 | | 2.3 | | 0.5 |  |
| 50-59 | 7.53 | | 3.3 | | 2.3 | | 2.3 |  |
| 60-74 | 7.02-7.93 | | 3.3 | | 2.3 | | 2.3 |  |
| 75-84 | 7.93 | | 3.3 | | 2.3 | | 2.3 |  |
| 85+ | 7.02 | | 3.3 | | 2.3 | | 2.3 |  |
| Percentage of vaccine doses with adverse event | | | | | | | | |
| Local grade 3 | 1.4% (0.0%, 1.4%) | | | | | | | Wilson et al. (2023) [16] |
| Systemic grade 3 | 1.1% (0.0%, 1.1%) | | | | | | |  |

CI = confidence interval; DSA = deterministic sensitivity analysis; RSV-ARD = RSV-related acute respiratory disease; RSV-LRTD = RSV-related lower respiratory tract disease; RSV = respiratory syncytial virus; US = United States.

*Values were not varied in sensitivity analyses.

^‡^ The proportions with RSV-LRTD and RSV-No LRTD sum to 100% in the model; accordingly, the proportions with RSV-No LRTD were calculated by subtracting the proportions with RSV-LRTD from 100%.

Table S-2. Deterministic Sensitivity Analysis Parameters

| Model parameter | Base-case value | Range applied in DSA |
| --- | --- | --- |
| RSV-ARD Incidence | 18-59: 6.7% | 5.4%; LB calculated as 80% of the mean |
|  |  | 8.0%; UB calculated as 120% of the mean |
|  | 60+ 5.7% | 4.6%; LB calculated as 80% of the mean |
|  |  | 6.9%; UB calculated as 120% of the mean |
| Percentage RSV-LRTD | 18-59: 15.2% | 12.2%; LB calculated as 80% of the mean |
|  |  | 18.2%: UB calculated as 120% of the mean |
|  | 60-64: 13.5% | 10.8%; LB calculated as 80% of the mean |
|  |  | 16.1%: UB calculated as 120% of the mean |
|  | 65-74: 14.5% | 11.6%; LB calculated as 80% of the mean |
|  |  | 17.2%: UB calculated as 120% of the mean |
|  | 75-84: 19.3% | 15.4%; LB calculated as 80% of the mean |
|  |  | 23.2%: UB calculated as 120% of the mean |
|  | 85+: 24.7% | 19.8%; LB calculated as 80% of the mean |
|  |  | 29.6%: UB calculated as 120% of the mean |
| Percentage with RSV-LRTD requiring hospitalization | 18-49: 7.1% | 5.7%; LB calculated as 80% of the mean |
|  |  | 8.5%; UB calculated as 120% of the mean |
|  | 50-59: 21.4% | 17.1%; LB calculated as 80% of the mean |
|  |  | 25.7%; UB calculated as 120% of the mean |
|  | 60-64: 13.7% | 11.0%; LB calculated as 80% of the mean |
|  |  | 16.5%: UB calculated as 120% of the mean |
|  | 65-74: 25.4% | 20.3%; LB calculated as 80% of the mean |
|  |  | 30.5%: UB calculated as 120% of the mean |
|  | 75-84: 28.9% | 23.1%; LB calculated as 80% of the mean |
|  |  | 34.7%: UB calculated as 120% of the mean |
|  | 85+: 31.2% | 24.9%; LB calculated as 80% of the mean |
|  |  | 37.4%: UB calculated as 120% of the mean |
| Percentage with RSV-LRTD requiring ED treatment | 18-49: 20.7% | 16.6%; LB calculated as 80% of the mean |
|  |  | 24.8%; UB calculated as 120% of the mean |
|  | 50-59: 17.4% | 13.9%; LB calculated as 80% of the mean |
|  |  | 20.8%; UB calculated as 120% of the mean |
|  | 60-64: 14.8% | 11.8%; LB calculated as 80% of the mean |
|  |  | 17.8%: UB calculated as 120% of the mean |
|  | 65-74: 20.1% | 16.1%; LB calculated as 80% of the mean |
|  |  | 24.1%: UB calculated as 120% of the mean |
|  | 75-84: 20.1% | 16.1%; LB calculated as 80% of the mean |
|  |  | 24.1%: UB calculated as 120% of the mean |
|  | 85+: 20.0% | 16.0%; LB calculated as 80% of the mean |
|  |  | 24.0%: UB calculated as 120% of the mean |
| Percentage with RSV-No LRTD requiring outpatient care | 18-49: 25.3% | 20.2%; LB calculated as 80% of the mean |
|  |  | 30.3%; UB calculated as 120% of the mean |
|  | 50-59: 29.9% | 23.9%; LB calculated as 80% of the mean |
|  |  | 35.9%; UB calculated as 120% of the mean |
|  | 60-64: 24.1% | 19.3%; LB calculated as 80% of the mean |
|  |  | 28.9%: UB calculated as 120% of the mean |
|  | 65-74: 33.4% | 26.8%; LB calculated as 80% of the mean |
|  |  | 40.1% UB calculated as 120% of the mean |
|  | 75-84: 39.6% | 31.7%; LB calculated as 80% of the mean |
|  |  | 47.6%: UB calculated as 120% of the mean |
|  | 85+: 47.3% | 37.9%; LB calculated as 80% of the mean |
|  |  | 56.8%: UB calculated as 120% of the mean |
| RSV-Related Mortality (among hospitalized cases) | 18-24: 2.4% | 1.9%; LB from Havers et al. (2024) |
|  |  | 6.0%; UB from Branche (2023) unpublished data presented at IDWeek |
|  | 25-49: 4.3% | 1.9%; LB from Havers et al. (2024) |
|  |  | 6.0%; UB from Branche (2023) unpublished data presented at IDWeek [5] |
|  | 50-59: 4.3% | 3.1%; LB from Havers et al. (2024) [6] |
|  |  | 6.0%; UB from Branche (2023) unpublished data presented at IDWeek[5] |
|  | 60-64: 7.6% | 3.9%; LB from Hutton et al. (2024)[7] |
|  |  | 7.6%; UB from Tseng et al. (2020)[8] |
|  | 65-74: 7.6% | 4.3%: LB from Hutton [7] |
|  |  | 7.6%: UB from Tseng et al. (2020)[8] |
|  | 75+: 7.6% | 5.7%: LB from Hutton [7] |
|  |  | 7.6%: UB from Tseng et al. (2020)[8] |
| RSV-LRTD Hospitalization Costs | $12,198 | $9,123; LB from Choi et al. (2022)[17] |
|  |  | $48.962; UB from Pastula et al. (2017)[4] |
| RSV ED Costs | 18-49: $3,258 | $2,334; LB from Wyffels [12]  $6,074: UB from Averin[18] |
|  | 50-74: $4,776 |  |
|  | 75-84: $4149 |  |
|  | 85+: $4379 |  |
| RSV Outpatient Costs | $2,334 | $128; LB from Hutton [19] |
|  |  | $2,334; UB assumed to be the same as base case[12] |
| RSV-LRTD & RSV-No LRTD QALY losses | Hospitalized: 0.0193 | 0.0154; LB calculated as 80% of the mean |
|  |  | 0.0232; UB calculated as 120% of the mean |
|  | Outpatient: 0.0185 | 0.0148; LB calculated as 80% of the mean |
|  |  | 0.0222; UB calculated as 120% of the mean |
|  | No treatment: 0.0093 (assumes to be half of outpatient) | 0.0074; LB calculated as 80% of the mean |
|  |  | 0111; UB calculated as 120% of the mean |
|  | ED: 0.0185 (assumes to be equal to outpatient) | 0.0148; LB calculated as 80% of the mean |
|  |  | 0.0222; UB calculated as 120% of the mean |
| Percent experiencing grade 3 adverse events | Grade 3 local: 1.43% | 0%; assume no AEs |
|  |  | 1.71%; UB calculated as 120% of the mean |
|  | Grade 3 systemic: 1.13% | 0%; assume no AEs |
|  |  | 1.36%; UB calculated as 120% of the mean |
| QALY lost due to grade 3 adverse events | Grade 3 local: 0.00027 | 0.00022; LB calculated as 80% of the mean |
|  |  | 0.00033; UB calculated as 120% of the mean |
|  | Grade 3 systemic: 0.0011 | 0.0009; LB calculated as 80% of the mean |
|  |  | 0.0013; UB calculated as 120% of the mean |
| Costs associated with grade 3 adverse events | Grade 3 local: $8 | $6; LB calculated as 80% of the mean |
|  |  | $9; UB calculated as 120% of the mean |
|  | Grade 3 systemic: $12 | $9; LB calculated as 80% of the mean |
|  |  | $14; UB calculated as 120% of the mean |
| Lifetime productivity loss due to premature RSV-related death (market value) | 18-24: $1,695,389 | $1,356,311; LB calculated as 80% of the mean |
|  |  | $2,034,467; UB calculated as 120% of the mean |
|  | 25-29: $1,744,274 | $1,395,419; LB calculated as 80% of the mean |
|  |  | $2,093,129; UB calculated as 120% of the mean |
|  | 30-34: $1,677,264 | $1,341,811; LB calculated as 80% of the mean |
|  |  | $2,012,716; UB calculated as 120% of the mean |
|  | 35-39: $1,540,571 | $1,232,457; LB calculated as 80% of the mean |
|  |  | $1,848,686; UB calculated as 120% of the mean |
|  | 40-44: $1,345,215 | $1,076,172; LB calculated as 80% of the mean |
|  |  | $1,614,258; UB calculated as 120% of the mean |
|  | 45-49: $1,104,447 | $883,558; LB calculated as 80% of the mean |
|  |  | $1,325,337; UB calculated as 120% of the mean |
|  | 50-54: $840,925 | $672,740; LB calculated as 80% of the mean |
|  |  | $1,009,110; UB calculated as 120% of the mean |
|  | 55-59: $572,470 | $457,976; LB calculated as 80% of the mean |
|  |  | $686,965; UB calculated as 120% of the mean |
|  | 60-64: $325,333 | $260,266; LB calculated as 80% of the mean |
|  |  | $390,399; UB calculated as 120% of the mean |
|  | 65-69: $155,192 | $124,153; LB calculated as 80% of the mean |
|  |  | $186,230; UB calculated as 120% of the mean |
|  | 70-74: $73,132 | $58,505; LB calculated as 80% of the mean |
|  |  | $87,758; UB calculated as 120% of the mean |
|  | 75-79: $33,758 | $27,007; LB calculated as 80% of the mean |
|  |  | $40,510; UB calculated as 120% of the mean |
|  | 80-84: $14,646 | $11,717; LB calculated as 80% of the mean |
|  |  | $17,576; UB calculated as 120% of the mean |
|  | 85+: $3,099 | $2,479; LB calculated as 80% of the mean |
|  |  | $3,718; UB calculated as 120% of the mean |
| Lifetime productivity loss due to premature RSV-related death (market value) | 18-24: $927,731 | $742,185; LB calculated as 80% of the mean |
|  |  | $1,113,277; UB calculated as 120% of the mean |
|  | 25-29: $930,977 | $744,782; LB calculated as 80% of the mean |
|  |  | $1,117,173; UB calculated as 120% of the mean |
|  | 30-34: $880,005 | $704,004; LB calculated as 80% of the mean |
|  |  | $1.056,006; UB calculated as 120% of the mean |
|  | 35-39: $783,997 | $627,198; LB calculated as 80% of the mean |
|  |  | $940,797; UB calculated as 120% of the mean |
|  | 40-44: $675,045 | $540,036; LB calculated as 80% of the mean |
|  |  | $810,054; UB calculated as 120% of the mean |
|  | 45-49: $579,194 | $463,356; LB calculated as 80% of the mean |
|  |  | $695,033; UB calculated as 120% of the mean |
|  | 50-54: $504,421 | $403,537; LB calculated as 80% of the mean |
|  |  | $605,305; UB calculated as 120% of the mean |
|  | 55-59: $444,594 | $355,676; LB calculated as 80% of the mean |
|  |  | $533,513; UB calculated as 120% of the mean |
|  | 60-64: $385,345 | $308,276; LB calculated as 80% of the mean |
|  |  | $462,414; UB calculated as 120% of the mean |
|  | 65-69: $313,119 | $250,495; LB calculated as 80% of the mean |
|  |  | $375,743; UB calculated as 120% of the mean |
|  | 70-74: $233,358 | $186,686; LB calculated as 80% of the mean |
|  |  | $280,030; UB calculated as 120% of the mean |
|  | 75-79: $154,041 | $123,232; LB calculated as 80% of the mean |
|  |  | $184,849; UB calculated as 120% of the mean |
|  | 80-84: $84,617 | $67,693; LB calculated as 80% of the mean |
|  |  | $101,540; UB calculated as 120% of the mean |
|  | 85+: $14,499 | $11,599; LB calculated as 80% of the mean |
|  |  | $17,399; UB calculated as 120% of the mean |

Table S-3. Detailed Results From Scenario Analyses by Age Group

|  | **mRNA-1345** | **No vaccination** | **Difference*** |
| --- | --- | --- | --- |
| **18-49 years, high risk** |  |  |  |
| Clinical outcomes (thousands of cases) |  |  |  |
| Hospitalizations | 119 | 140 | -21 |
| ED visits | 346 | 407 | -62 |
| Outpatient visits | 3,610 | 4,204 | -594 |
| Deaths | 3 | 4 | -.57 |
| QALYs lost | 6,089 | 6,122 | -32 |
| Economic outcomes ($ billions) |  |  |  |
| Direct medical costs | $10.4 | $12.1 | -$1.8 |
| Vaccination | $4.6 | $0.00 | $4.6 |
| Indirect costs | $14.0 | $16.3 | -$2.3 |
| Total costs (direct + vaccination + indirect) | $29.0 | $28.4 | $0.6 |
| ICER (Cost per QALY gained) |  |  | $18,027 |
| 50-59 years, high risk |  |  |  |
| Clinical outcomes (thousands of cases) |  |  |  |
| Hospitalizations | 134 | 167 | −34 |
| ED visits | 109 | 136 | −27 |
| Outpatient visits | 1,455 | 1,785 | −330 |
| Deaths | 6 | 7 | −1 |
| QALYs | 6,468 | 6,499 | −31 |
| Economic outcomes ($ billions) |  |  |  |
| Direct medical costs | $5.2 | $6.5 | -$1.3 |
| Vaccination | $2.4 | $0.0 | $0.2 |
| Indirect costs | $10.4 | $12.8 | $0.2 |
| Total costs (direct + vaccination + indirect) | $18.0 | $19.3 | -$0.1 |
| ICER (Cost per QALY gained) |  |  | Cost saving |
| **60-74 years, high risk** |  |  |  |
| Clinical outcomes (thousands of cases) |  |  |  |
| Hospitalizations | 316 | 428 | −111 |
| ED visits | 216 | 291 | −75 |
| Outpatient visits | 2,697 | 3,519 | −822 |
| Deaths | 24 | 32 | −8 |
| QALYs | 21,021 | 21,125 | −104 |
| Economic outcomes |  |  |  |
| Direct medical costs | $10.5 | $14.0 | -$3.5 |
| Vaccination | $5.7 | $0.0 | $5.7 |
| Indirect costs | $17.8 | $23.3 | -$5.6 |
| Total costs (direct + vaccination + indirect) | $3.4 | $37.3 | -$3.4 |
| ICER (Cost per QALY gained) |  |  | Cost saving |
| **75+ years, all** |  |  |  |
| Clinical outcomes (thousands of cases) |  |  |  |
| Hospitalizations | 267 | 374 | −107 |
| ED visits | 182 | 255 | −73 |
| Outpatient visits | 1,976 | 2,671 | −694 |
| Deaths | 20 | 28 | −8 |
| QALYs | 31,117 | 31,171 | −54 |
| Economic outcomes ($ billions) |  |  |  |
| Direct medical costs | $8.1 | $11.2 | -$3.1 |
| Vaccination | $5.3 | $0.0 | $5.3 |
| Indirect costs | $4.3 | $5.9 | -$1.6 |
| Total costs (direct + vaccination + indirect) | $17.8 | $17.1 | $0.6 |
| ICER (Cost per QALY gained) |  |  | 11,714 |
| **60+ years, all** |  |  |  |
| Clinical outcomes (thousands of cases) |  |  |  |
| Hospitalizations | 609 | 836 | −227 |
| ED visits | 477 | 652 | −174 |
| Outpatient visits | 6,056 | 7,980 | −1,925 |
| Deaths | 46 | 63 | −17 |
| QALYs | 71,291 | 71,471 | −180 |
| Economic outcomes ($ billions) |  |  |  |
| Direct medical costs | $22.3 | $30.0 | -$7.7 |
| Vaccination | $16.1 | $0.0 | $16.2 |
| Indirect costs | $27.6 | $36.2 | -$8.6 |
| Total costs (direct + vaccination + indirect) | $66.1 | $66.2 | -$0.1 |
| ICER (Cost per QALY gained) |  |  | Cost saving |

ED = emergency department; ICER = incremental cost-effectiveness ratio; QALY = quality-adjusted life-year.

*mRNA-1345 versus No vaccination

Table S-4. Detailed Economic Outcomes from Scenario Analyses Considering a Healthcare Perspective and Excluding Caregiver Losses

|  | mRNA-1345 | | No vaccination | Difference* |
| --- | --- | --- | --- | --- |
| **Healthcare perspective** | | | |  |
| Economic outcomes (% billions) | | | |  |
| Direct medical costs | $37.9 | | $48.7 | −$10.7 |
| Vaccination | $23.2 | | $0.0 | $23.2 |
| Indirect costs | $52.0 | | $65.3 | −$13.3 |
| Total costs (direct + vaccination + indirect) | $61.2 | | $48.7 | $12.5 |
| ICER (Cost per QALY gained) |  |  | | $51,200 |
| **No caregiver** | | | |  |
| Economic outcomes ($ billions) | | | |  |
| Direct medical costs | $37.9 | | $48.7 | −$10.7 |
| Vaccination | $23.2 | | $0.0 | $23.2 |
| Indirect costs | $52.0 | | $65.3 | −$12.9 |
| Total costs (direct + vaccination + indirect) | $61.2 | | $48.7 | −$0.5 |
| ICER (Cost per QALY gained) |  | |  | Cost saving |

Note: Clinical outcomes are the same as the base case

ED = emergency department; ICER = incremental cost-effectiveness ratio; QALY = quality-adjusted life-year.

*mRNA-1345 versus No vaccination

Table S-5. Detailed Results from Scenario Analyses Considering Linear Waning

|  | mRNA-1345 | | No vaccination | Difference* | |
| --- | --- | --- | --- | --- | --- |
| **2-year time horizon** | | | |  | |
| Clinical outcomes (thousands of cases) | | | |  | |
| Hospitalizations | 280 | | 479 | −199 | |
| ED visits | 310 | | 494 | −184 | |
| Outpatient visits | 4,113 | | 5,757 | −1,644 | |
| Deaths | 18 | | 31 | −14 | |
| QALYs lost | 35,301 | | 35,466 | −164 | |
| Economic outcomes ($ billions) | | | |  | |
| Direct medical costs | $14.1 | | $21.0 | −$7.0 | |
| Vaccination | $23.2 | | $0.0 | $23.2 | |
| Indirect costs | $19.2 | | $27.8 | −$8.6 | |
| Total costs (direct + vaccination + indirect) | $56.5 | | $48.9 | $7.7 | |
| ICER (Cost per QALY gained) |  |  | | | $46,535 |
| **3-year time horizon** | | | |  | |
| Clinical outcomes (thousands of cases) | | | |  | |
| Hospitalizations | 462 | | 707 | −245 | |
| ED visits | 505 | | 732 | −227 | |
| Outpatient visits | 6,618 | | 8,548 | −1,930 | |
| Deaths | 29 | | 46 | −17 | |
| QALYs | 52,039 | | 52,237 | −197 | |
| Economic outcomes | | | |  | |
| Direct medical costs | $22.4 | | $30.7 | −$8.3 | |
| Vaccination | $23.2 | | $0.0 | $23.2 | |
| Indirect costs | $30.4 | | $40.9 | −$10.5 | |
| Total costs (direct + vaccination + indirect) | $76.0 | | $71.6 | $4.4 | |
| ICER (Cost per QALY gained) |  | |  | $22,467 | |

ED = emergency department; ICER = incremental cost-effectiveness ratio; QALY = quality-adjusted life-year.

*mRNA-1345 – no vaccination.

Table S-6. Detailed Results from Scenario Analyses with Individual Risk Conditions (18-59 HR, 5-year time horizon, societal perspective)

|  | mRNA-1345 | No vaccination | Difference* |
| --- | --- | --- | --- |
| **Asthma** |  |  |  |
| Clinical outcomes (thousands of cases) |  |  |  |
| Hospitalizations | 57 | 69 | -12 |
| ED visits | 91 | 109 | -18 |
| Outpatient visits | 2,310 | 2,718 | -408 |
| Deaths | 2 | 2 | -0.5 |
| QALYs lost | 5,982 | 6,005 | -23 |
| Economic outcomes ($ billions) |  |  |  |
| Direct medical costs | $6.0 | $7.2 | -$1.1 |
| Vaccination | $3.6 | $0.0 | $3.6 |
| Indirect costs | $8.1 | $9.6 | -$1.5 |
| Total costs (direct + vaccination + indirect) | $17.8 | $16.8 | $1.0 |
| ICER (Cost per QALY gained) |  |  | $43,418 |
| **Diabetes (with and without complications)** |  |  |  |
| Clinical outcomes (cases) |  |  |  |
| Hospitalizations | 50 | 62 | -12 |
| ED visits | 70 | 84 | -15 |
| Outpatient visits | 1,017 | 1,214 | -197 |
| Deaths | 2 | 2 | -0.5 |
| QALYs | 3,196 | 3,210 | -14 |
| Economic outcomes |  |  |  |
| Direct medical costs | $3.1 | $3.7 | -$0.6 |
| Vaccination | $1.6 | $0.0 | $1.6 |
| Indirect costs | $5.0 | $6.0 | -$1.0 |
| Total costs (direct + vaccination + indirect) | $9.6 | $9.7 | -$0.1 |
| ICER (Cost per QALY gained) |  |  | Cost saving |
| **Coronary artery disease (CAD)** |  | |  |
| Clinical outcomes (thousands of cases) |  | |  |
| Hospitalizations | 14 | 18 | -3 |
| ED visits | 14 | 17 | -3 |
| Outpatient visits | 183 | 222 | -39 |
| Deaths | 0.6 | 0.8 | -0.1 |
| QALYs | 687 | 690 | -3 |
| Economic outcomes ($ billions) |  |  |  |
| Direct medical costs | $0.6 | $0.8 | -$0.1 |
| Vaccination | $0.3 | $0.0 | $0.3 |
| Indirect costs | $1.2 | $1.4 | -$0.3 |
| Total costs (direct + vaccination + indirect) | $2.1 | $2.2 | -$0.1 |
| ICER (Cost per QALY gained) |  |  | Cost-saving |
| **Chronic kidney disease (all ages)** |  |  |  |
| Clinical outcomes (cases) |  |  |  |
| Hospitalizations | 13 | 16 | -3 |
| ED visits | 14 | 17 | -3 |
| Outpatient visits | 175 | 211 | -36 |
| Deaths | 0.5 | 0.6 | -0.1 |
| QALYs | 617 | 620 | -3 |
| Economic outcomes |  |  |  |
| Direct medical costs | $0.6 | $0.7 | -$0.1 |
| Vaccination | $0.3 | $0.0 | $0.3 |
| Indirect costs | $1.1 | $1.3 | -$0.2 |
| Total costs (direct + vaccination + indirect) | $1.9 | $2.0 | -$0.1 |
| ICER (Cost per QALY gained) |  |  | Cost saving |
| **Chronic liver disease** |  |  |  |
| Clinical outcomes (thousands of cases) |  |  |  |
| Hospitalizations | 20 | 25 | -4 |
| ED visits | 30 | 36 | -6 |
| Outpatient visits | 348 | 414 | -66 |
| Deaths | 0.7 | 0.9 | -0.1 |
| QALYs lost | 988 | 993 | -5 |
| Economic outcomes ($ billions) |  |  |  |
| Direct medical costs | $1.1 | $1.3 | -$0.2 |
| Vaccination | $0.5 | $0.0 | $0.5 |
| Indirect costs | $1.8 | $2.2 | -$0.4 |
| Total costs (direct + vaccination + indirect) | $3.4 | $3.5 | -$0.1 |
| ICER (Cost per QALY gained) |  |  | Cost saving |
| **Chronic obstructive pulmonary disease (COPD)** |  |  |  |
| Clinical outcomes (thousands of cases) |  |  |  |
| Hospitalizations | 28 | 35 | -7 |
| ED visits | 27 | 34 | -6 |
| Outpatient visits | 299 | 362 | 63 |
| Deaths | 1 | 1 | 0.3 |
| QALYs | 1,064 | 1,071 | 6 |
| Economic outcomes ($ billions) |  |  |  |
| Direct medical costs | $1.1 | $1.3 | -$0.2 |
| Vaccination | $0.5 | $0.0 | $0.5 |
| Indirect costs | $2.1 | $2.6 | -$0.5 |
| Total costs (direct + vaccination + indirect) | $3.7 | $3.9 | -$0.3 |
| ICER (Cost per QALY gained) |  |  | Cost saving |
| **Congestive heart failure** |  |  |  |
| Clinical outcomes (thousands of cases) |  |  |  |
| Hospitalizations | 24 | 30 | -6 |
| ED visits | 30 | 36 | -6 |
| Outpatient visits | 182 | 218 | -37 |
| Deaths | 1 | 1 | -0.2 |
| QALYs | 494 | 498 | -4 |
| Economic outcomes (% billion) |  |  |  |
| Direct medical costs | $0.8 | $1.0 | -$0.1 |
| Vaccination | $0.2 | $0.0 | $0.2 |
| Indirect costs | $1.6 | $2.0 | -$0.4 |
| Total costs (direct + vaccination + indirect) | $2.6 | $2.9 | -$0.3 |
| ICER (Cost per QALY gained) |  |  | Cost saving |
| **Morbid obesity (BMI>40)** |  |  |  |
| Clinical outcomes (thousands of cases) |  |  |  |
| Hospitalizations | 81 | 99 | -17 |
| ED visits | 159 | 189 | -31 |
| Outpatient visits | 1,746 | 2,058 | -312 |
| Deaths | 3 | 3 | -0.6 |
| QALYs | 4,057 | 4,078 | -21 |
| Economic outcomes |  |  |  |
| Direct medical costs | $5.3 | $6.3 | -$1.0 |
| Vaccination | $2.4 | $0.0 | $2.4 |
| Indirect costs | $8.1 | $9.6 | -$1.5 |
| Total costs (direct + vaccination + indirect) | $15.8 | $15.9 | -$0.1 |
| ICER (Cost per QALY gained) |  |  | Cost saving |
